# Understanding the barriers and facilitators to using self-sampling packs for sexually transmitted infections and blood borne viruses: thematic analyses supporting intervention optimisation

**DOI:** 10.1101/2020.11.22.20230359

**Authors:** Paul Flowers, Maria Pothoulaki, Gabriele Vojt, Fiona Mapp, Melvina Woode Owusu, Claudia Estcourt, Jackie Cassell, John Saunders

## Abstract

**Purpose:** This paper illustrates initial steps of an intervention optimisation process. Self-sampling packs for sexually transmitted infections (STIs) and blood borne viruses (BBVs) are widely offered within the UK, yet have problems with reach and sample return rates. They have arisen without any formal intervention development.

**Methods:** Eleven focus groups and seven interviews were conducted with convenience samples of patients recruited from sexual health clinics and members of the public in late 2017 (n=57). To enable intervention optimisation firstly we formulated initial programme theory situating the intervention. Secondly, we conducted an inductive appraisal of the behavioural system of using the pack to understand meaningful constituent behavioural domains. Subsequently we conducted a thematic analysis of barriers and facilitators to enacting each sequential behavioural domain in preparation for future behaviour change wheel analysis.

**Results:** Overall, we found that self-sampling packs were acceptable. Our participants understood their overall logic and value as a pragmatic intervention that simultaneously reduced barriers to, and facilitated, individuals being tested for STIs. However, at the level of each behavioural domain (e.g., reading leaflets, returning samples), problems with the pack were identified as well as a series of potential optimisations which might widen the reach of self-sampling and increase the return of viable samples.

**Conclusions:** This paper provides an example of a pragmatic approach to optimising an intervention already widely offered across the UK. The paper demonstrates the added value health psychological approaches make; systematically considering the context of the intervention, in addition to illuminating granular areas for improvement.

**What is already known on this subject?:**

- The use of self-sampling packs for sexually transmitted infections (STIs) and blood borne viruses (BBVs) has been widely implemented without in-depth assessment of user engagement or theorisation
- Some evidence suggests that the uptake of self-sampling packs, and the concomitant return of samples to enable diagnosis, are socially patterned
- Despite increasing and widespread use of self-sampling packs across the UK, relatively little is currently known about their acceptability, or how they could be improved

**What does this study add?:**

- This study provides an illustrative example of using a preliminary programme theory to situate the problem to be addressed by intervention optimisation
- The thematic analyses show that self-sampling packs offer a largely acceptable means to enabling STI and BBV testing and diagnosis; they remove many barriers to testing. However, several modifiable barriers to use endure, potentially reducing sample return and amplifying health inequalities
- This study presents a range of barriers and facilitators to the various behavioural domains included within the use of self-sampling packs. It summarises the findings ready for subsequent behaviour change wheel analyses

## Introduction

The World Health Organization (WHO) has estimated a global burden of 376 million newly diagnosed sexually transmitted infections (STIs) per year (World Health Organization, 2018). The WHO highlights an overall lack of progress in preventing STI transmission over time. Care of people with STIs and blood borne viruses (BBVs) is complex and improving outcomes is challenging; infections are often asymptomatic. STIs and BBVs disproportionately affect some people more than others, are stigmatised, and are associated with typical health inequalities such as sexual identity, deprivation, non-white ethnicity and geographic factors (Woodhall *et al*., 2016; Tanton *et al*., 2017). In response to increased disease burden and reduced funding for STI care and prevention, novel models of sexual healthcare delivery have rapidly emerged without an evidence base.

One such model is the provision of home delivered self-sampling packs for STIs and BBVs. These packs are used by a person requesting a kit online, taking their own samples at home with a series of kits, sending the completed samples to a laboratory for testing, and usually receiving results by text message. Given the recent impact of COVID-19 it seems highly likely that this kind of home-based self-sampling and testing will become more commonplace across a range of health domains. Health psychology has much to offer in assisting with this intervention development and implementation.

In relation to the current study we developed an initial programme theory (Figure 1) as a conceptual tool to assist with the process of intervention optimisation (for more information on programme theory, see MRC guidance (Medical Research Council, due 2020), or individual papers such as Brousselle and Champagne (2011) and Rogers (2008)). It shows how these packs are thought to work at multiple levels for users, providers, and other stakeholders such as commissioners. The context in which self-sampling packs have developed relates to wider societal changes in relation to self-managed health care, the shift to eHealth, the postal delivery of a broad range of commodities (from groceries, to books and furniture), and changes in patterns of sexual mixing that are associated with increased transmission of STIs and BBVs (e.g. dating apps facilitate increased disassortative sexual mixing).

**Figure 1:**
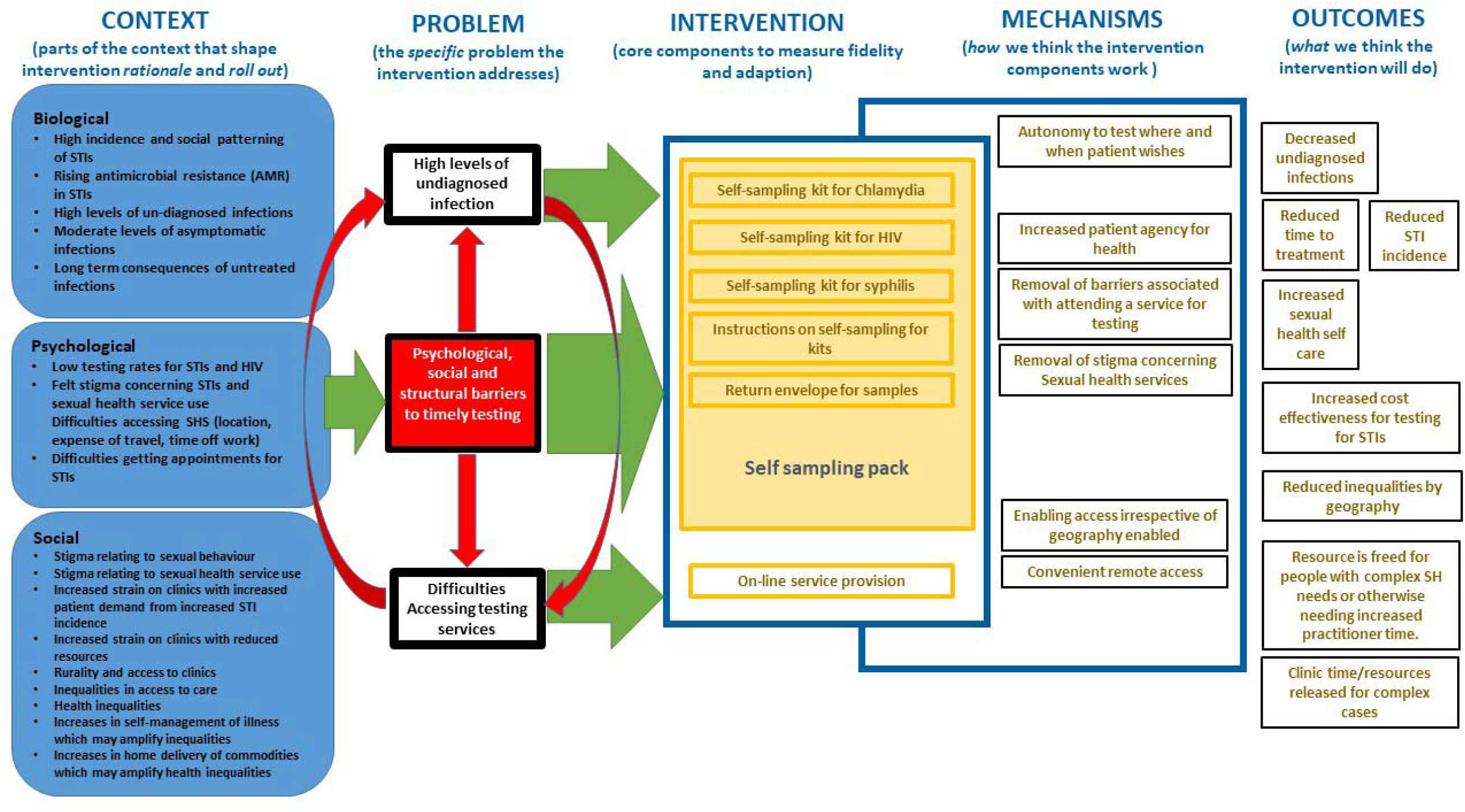
A preliminary programme theory detailing how self-sampling packs work.

Drawing on the biopsychosocial model (Engel, 1977), our preliminary programme theory (Figure 1) illustrates the ways that self-sampling packs provide a simple solution that simultaneously addresses multiple problems, pitched at varied biological, psychological and social levels (illustrated on the left-hand side of the figure). These problems represent the current ‘perfect storm’ (White, 2017), of changing epidemiology, changing norms and a crisis in service provision. These are set against a background of on-going stigma shaping a range of sexual health related behaviours (e.g. Morris et al., 2014). The programme theory shows how the logic of self-sampling packs works well for pack users, for health care professionals, and more broadly, for the health care systems and for public health.

In this way, it is not surprising that self-sampling for STIs has been widely and rapidly adopted throughout the UK as a pragmatic response to these multiple drivers across the health care system and wider communities of users. In some areas of England it is intended to be the primary means to enabling asymptomatic patients to get tested and diagnosed (Campbell and Marsh, 2017), and this may be beneficial to many people who cannot easily access health facilities in person. However, evidence is emerging of unanticipated adverse consequences. Studies suggest that self-sampling is under used by older people, by men, by non-heterosexual women, by black, minority and ethnic populations (Kersaudy-Rahib *et al*., 2017; Manavi and Hodson, 2017; Banerjee *et al*., 2018; Barnard *et al*., 2018). Moreover, sample return rates range considerably, for example 54% in the National HIV Self-sampling service (Guerra et al., 2016) compared to 72.5% of test kits returned to SH:24 covering two London boroughs (Barnard *et al*., 2018). Moreover, treatment rates are lower in individuals using self-sampling than those attending sexual health services (Banerjee et al., 2018; Kersaudy-Rahib *et al*., 2017; Manavi & Hodson, 2017).

In this paper, we report the first step of a multi-staged intervention development process which sought to optimise the use of self-sampling packs. The analysis reported here was followed by a behaviour change wheel (Michie et al., 2011) analysis to specify potential intervention components that could enable optimisation (Flowers et al., forthcoming 2020). Although there is a growing literature regarding the process of intervention development *per se*, the equally important process of intervention optimisation is less well understood. While many key processes, and the use of health psychological tools and acumen are the same, divergence lies in considering the goals of the overall process. When *developing* an intervention, major practical concerns typically relate to acceptability, practicability and future implementability. These should be central to the overall process. In contrast, when *optimising* an existing intervention, broad brush issues of acceptability and implementability should already have been established. As a consequence, given the intervention is already up and running, there may be a more distinct focus on optimising the pre-existing intervention with new behaviour change content to extend the range of access and population reach to those most in need, to reduce inequalities and address emerging unintended negative intervention effects. From a methodological perspective intervention development typically focuses on ‘behavioural diagnosis’ in which, within a given behavioural system, a single key fulcrum point for investing in behavioural analysis is chosen (Michie et al., 2014). By contrast in intervention optimisation, a more holistic approach to understanding the behavioural system may be taken with a view to exploring and understanding how particular behaviours and their associated behavioural domains are connected. Where the interdependent behavioural elements have not already been set out in intervention development, these need to form the focus of optimisation in order to ensure plausible points of optimisation, as in this case.

## Methods

### Participants

Eleven focus groups and seven interviews with young heterosexual individuals and men who have sex with men (MSM) were conducted in Glasgow and London in late 2017. MSM and heterosexuals did not take part in the same focus groups. In total 57 participants took part in the study, of whom 23 were female and 34 were male. All were cisgender. 56% of the sample (n=32) were between 18 and 25 years of age. Most participants were of a ‘white British’ or ‘other white’ ethnic group (n=44) and reported ‘University’ as their highest level of education (n=38). Just over half were heterosexual (n=30) with a substantial minority of MSM. Participants included sexual health clinic attenders diagnosed with an STI (including HIV) in the past six months, and other members of the general public. Inclusion criteria for the latter were: ability to give informed consent and age (heterosexual participants aged 18-30 years and MSM 18-65 years). Some, but not all, of these participants had previously had an STI.

### Procedure

Healthcare providers offered clinic attenders the opportunity to take part in the study in three NHS Trusts and Health Boards in England and Scotland. Other members of the public were recruited through purposive and convenience sampling, including snowballing, advertising on social media and through the team’s personal social networks. Posters, formal letters and/or emails were used to approach stakeholder organisations and diverse work settings to further widen recruitment.

Data were collected within the focus groups and interviews using a topic guide and an example of a self-sampling and treatment (APT) pack. The pack contained antibiotics, condoms, information leaflets about chlamydia, gonorrhoea and HIV and syphilis and how to take a sample, chlamydia and gonorrhoea self-sample kit, HIV and syphilis self-sample kit, envelope for return of self-sample kits, request form for the sample to be processed by the lab and APT pack packaging (envelope or small box, no branding or other identifiable markings, and which fits through standard letterboxes). It is used within the context of accelerated partner therapy (APT), a form of partner notification intervention. Partner notification (PN) is the process of identifying, testing and treating sex partners of a person with a sexually transmitted infection (STI) (Cowan et al., 1996). APT is a method to speed up the enhanced patient referral process. In brief, the healthcare professional performs a telephone consultation with the sex partner in private during the index patient’s clinic attendance. If medically safe, the index patient receives an APT pack, containing antibiotics and self-sampling kits for STI and HIV to deliver to their sex partner(s), or the clinic can post the APT pack to the sex partner (Estcourt *et al*., 2020). In pilot studies, APT resulted in faster sex partner treatment and greater overall numbers of sex partners treated, when compared with routine care but lower levels of testing for HIV and other STIs, when offered without HIV testing as part of the pack (Estcourt *et al*., 2012, 2015). In this way, pertinent to the work presented here, the pack also included antibiotic treatment.

A topic guide was used to steer discussion across the full range of issues detailed; where possible within the focus groups discussion between participants was encouraged rather than between facilitators and participants. In this way the data collected were primarily concerned with understanding the barriers and facilitators to the implementation of each step of APT. For the analysis presented within this paper however, we focus only upon the data that related to the use of home-based self-sampling packs. The ways in which we addressed other key steps of APT are addressed elsewhere (Pothoulaki *et al*., in preparation)

Data were transcribed, anonymised and imported into NVivo (Version 10) (Computer Software for Qualitative Data Analysis). Initially one analyst (PF) led an inductive analysis exploring the data in relation to the behavioural system and associated behavioural domains associated with the use of the self-sampling packs. Subsequently, deductive analysis focussed *a priori* on the barriers and facilitators to each of the sequential steps identified. In this way, the analysis balanced an inductive understanding of how participants understood the distinct behavioural elements of the self-sampling pack (e.g., system vs domain vs specific behaviours) and a deductive analysis of the barriers and facilitators to each of these distinct steps. One analyst (PF) then led the deductive analysis of barriers and facilitators to each step. Two of the researchers who collected data (MP and GV) then audited this analysis. Disagreements were resolved through discussion. Where possible and appropriate the data are presented as interactive exchanges illustrating the social nature of the focus groups. The selected extracts are representative of the underlying theme reflecting the participants’ perspectives.

### Research questions

1. What are the meaningful behavioural domains of using self-sampling packs for sexually transmitted infections?
2. What are barriers and facilitators to enacting each of the behavioural domains?

## Results

### Research question 1. What are the meaningful behavioural domains of using self-sampling packs for sexually transmitted infections?

Our inductive analysis highlighted four key behavioural domains that appeared meaningful to the participants. These are illustrated within Figure 2. Each of the four sequential steps represents a distinct set of highly related specific behaviours, each addressing a coherent behavioural challenge that future behaviour change intervention content could address. Together these domains and concomitant individual behaviours can be thought of as representing the behavioural system of self-sampling pack use.

**Figure 2:**
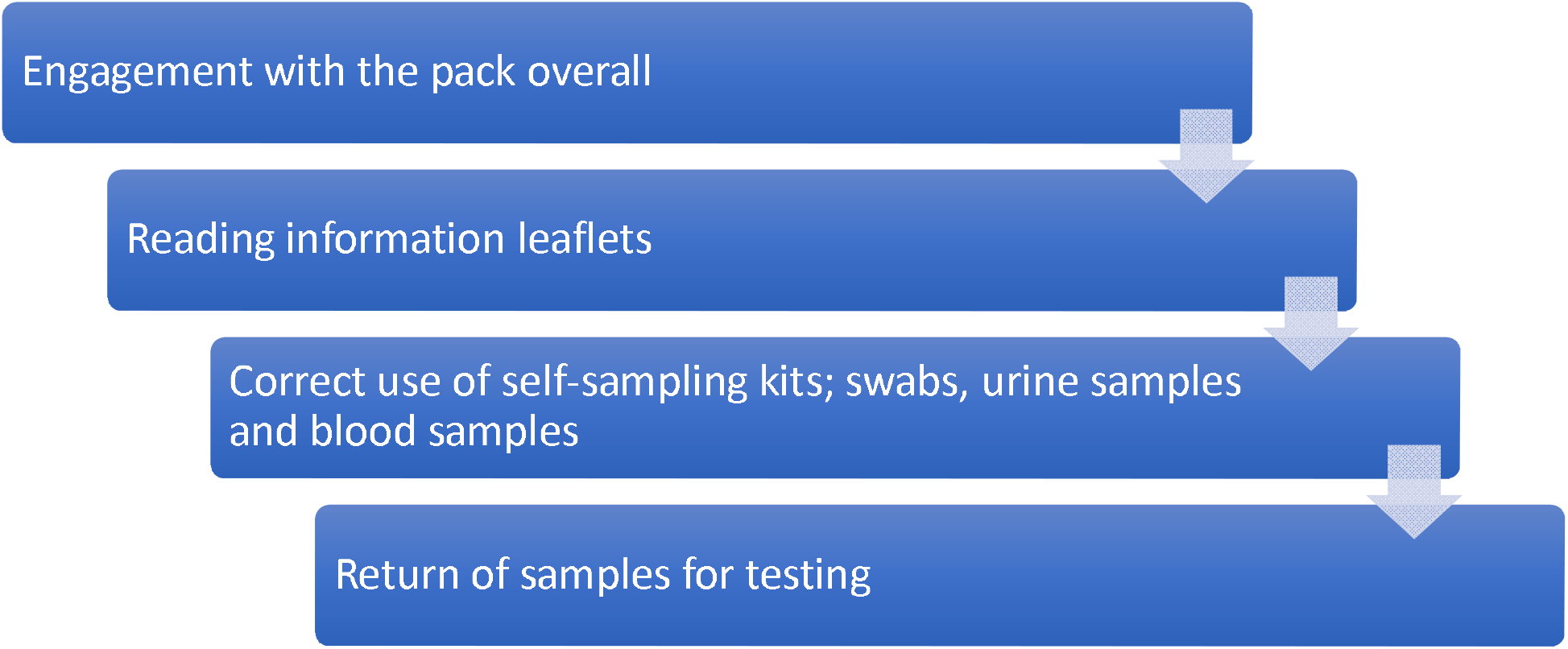
Inductively derived themes showing participants’ perspectives of the sequential behavioural domains of using self-sampling packs.

### Research question 2. What are barriers and facilitators to enacting each of the behavioural domains?

In relation to the second research question, results are structured according to the four key sequential steps associated with the overall behavioural system of using the self-sampling pack.

#### Pack users engaging with the pack overall

The pack, as a single entity, shapes the pack user’s subsequent engagement with the overall pack contents and their use of the kits within it. As suggested in Figure 2, participants (P) understood and clearly valued the logic of the self-sampling pack. For example:

> P1: I think the pack is very convenient, [… .] I think anybody would feel very uncomfortable going into the clinic and getting it done, reason why I say it’s convenient because you can do it in your own space, not to say that nobody’s up there to harm you or laugh at you doing anything but it’s just much more comfortable and convenient for the person themselves because it is quite embarrassing, I mean personally speaking it is very embarrassing but…
>
> Facilitator 1: Do you mean going to the clinic?
>
> P1: Going into the clinic and, you know, especially if you know what you’re going in for, so, hence why I think it’s, I think it’s actually incredible to be honest. I think it’s very, very convenient, and yeah.
>
> (Heterosexual women, patients, London, focus group)

However, beyond general appreciation for the logic of the overall pack, participants almost universally identified a series of barriers to engagement with the pack. There was consensus regarding the pack overall being confusing to potential users. Participants talked about being overwhelmed when they first looked at it: *‘It just exploded in for me and I don’t really know where to start’* (Heterosexual man, public, Glasgow, focus group). Participants also suggested ways of ameliorating this, clearly stating that there should be guidance explaining the structure and contents of the pack. These overall ‘pack-level’ instructions should include clear visuals and simple messages highlighting exactly what to do with the pack and its varied contents and the order in which to do them.

Participants believed that the organisation and structure of the pack were also important as it had a potential role in reducing pack users’ stress levels and enabling them to comply with specific instructions for the self-sampling kits within it. Participants recognised that pack users’ engagement with the pack may well take place in stressful circumstances; particularly when this occurs in the context of partner notification, or any other time when there is heightened perceived likelihood of infection. Pack use was likely to be stressful and high stress levels could negatively shape attention and decision-making processes relating to self-sampling kit use. Adding to the strong feelings about pack design, participants also raised further issues about the life circumstances of some pack users which could impact upon pack use. Participants outlined the ways such circumstances might amplify some of the problematic elements of pack design. The potential age of pack users, their disabilities, or domestic circumstances were understood to be important in relation to using the pack *‘It’s a mess, if you live in shared accommodation or with family and you want to keep it secret or something, it would be hard to stuff all this away, if Mum comes upstairs* (Heterosexual men, public, London, focus group). Participants also suggested the contents of the kit could be presented within a clearer physical framework, variously referred to as an ‘airline meal’, ‘gadget packaging’, ‘a chocolate box’ or a ‘graze box’ ™ in which individual components are packed within a stable card/plastic structure which allows each one to be seen. This attention to *how* to present the contents of the pack overall did not relate to aesthetics but to pack usability. A well-designed pack could enhance pack user self-efficacy, minimise perceived risks of non-compliance and, for a minority, indicate the degree of professional *care* in the absence of a face to face interaction. Participants also suggested the potential of having online video support that could not only assist with the use of the pack overall but also provide in-depth information about chlamydia and its health consequences.

#### Pack users reading and engaging with the information leaflets

Participants also outlined several barriers and facilitators to reading the leaflets within the pack itself. They highlighted the importance of literacy and health literacy levels and ability to retain attention as well as highlighting the particular challenges of engaging with the information leaflets if English was not a first language or if pack users were dyslexic or had learning difficulties. The extract below, for example, shows how important the leaflets could be in shaping pack users’ understandings of the consequences of the infection. In turn this could be important for wider engagement with treatment and self-sampling:

> Facilitator 2: You said earlier on that it was really important that people actually understood what sexually transmitted infections can do to the body.
>
> P1: Aye … the information sheet?
>
> Facilitator 2: Aye.
>
> P1: This is the same one I got shown.
>
> Facilitator 2: So, see as part of the pack that’s exactly what you get, do you think that’s, I mean do you find that useful?
>
> P1: Sometimes it can, looking at it, it could be too much for somebody to read. Just break it down into detail, like wee [small], short sentences that physically somebody could understand. Whereas, you might get somebody that will just read halfway through it, and just go, ‘Oh do you know what… … ‘Get that away from me!’. [… …] Think about it… if you shorten it down to two pages, short and sweet.
>
> Facilitator 2: So, essentially re-structure this, make it shorter and I think you said just keep it to the essentials, make it easier to read?
>
> P1: Aye.
>
> (Heterosexual men, patients, Glasgow, focus group)

Participants agreed that breaking the text up into smaller units and the use of better visuals may enable the pack users to better engage with self-sampling kits:

> P1: Aha, yeah. Like a small thing and it’s all in little paragraphs kind of thing. I think that those little leaflets are really easy to read, and I’m quite happy to just sit and read one of them in my own time. Whereas this kind of looks like a bank statement to me a little bit and I’m a little bit like… no thank you. It just looks like something that my mum and dad would get in the post, not something that I would sit and read, if that makes sense. Facilitator 2: What’s the difference between your mum and dad and you? Can you be a bit more specific?
>
> P1: Well, it looks really formal, like something that you would receive, like a bill or something in the post. Just the way it’s all laid out.
>
> (Heterosexual women, patients, Glasgow, focus group)

Participants also highlighted the potential of improving the leaflets further with more detail regarding how long the self-sampling process would take to complete.

#### Correct use of self-sampling kits; swabs, urine samples and blood samples

For many of the participants, the self-sampling kits within the pack were seen as relatively unproblematic and straightforward. However, one participant within one focus group struggled with the idea of self-sampling. For her, a key barrier to self-sampling related to how she perceived the professional roles and responsibilities of clinical staff *‘Why am I doing the doctor’s job? Why am I taking my own test? I’m sorry, I’m pure getting annoyed if you’re telling me to do my own… sorry. […] What if I do it wrong and then it comes back inconclusive and then there’s… wasted even more money. Or, like, goes back saying that I don’t have it when actually I do have it’* (Heterosexual women, public, London, focus group). Such worries over the accuracy of the test results that came from self-sampling were expressed by several participants across focus groups. More broadly, engagement with the pack’s multiple self-sampling kits, for a variety of STIs was seen by some to be an opportunity to check wider sexual health in a convenient and time efficient manner. Perhaps the biggest barrier to complying with the various self-sampling kits was the inclusion of the HIV self-sampling kit within the pack. This was understood to be problematic for a few reasons; detracting from the focus upon bacterial STIs; the particular stigma associated with HIV; the perceived gravity of an HIV positive status; challenges with blood-based sample collection such as fear of needles, or perceptions of not being able to collect a viable sample of blood.

> P3: And I suppose the emotional stress as well that you’re going through, or that I would be going through, thinking, have I potentially got an STI? And then trying to do a pack like that at home, I would probably find that quite stressful. Like, I’d already be stressed and then I’d be like, ‘How the hell do you use this fucking thing?’ And then I’d be like, ‘Bastard!’ [voices overlap]…
>
> [General laughter].
>
> P3: It stressed me out [voices overlap 48:56]
>
> P1: Just… can I just add as well, one minute he had chlamydia and then… I think it was pure, like, ‘Aye! We’re going to test you for HIV!’. I don’t know about anybody else, but if that was me, I’d be like, ‘What the fuck are you talking about HIV? I’d be like, I thought I had chlamydia. I know, like, you do that ‘cause if you’re pregnant, they do give that test and all that as well, like… what is it, like, hep C and all that…
>
> P2: Hepatitis.
>
> P1: Aye, ‘cause I even… I knew I didn’t have it, but when I was waiting on those results, I was like, I hope I don’t have it. I hope I don’t have it. I just… it’s one of those things, know what I mean. Imagine hearing that on the phone. You’d be like, ‘Why are you testing me for HIV? I thought it was just chlamydia.’ P3: So maybe just focus on, like, the problem in hand…
>
> P1: The problem in hand, yeah.
>
> P3: … and not necessarily talking about other bigger things if you were already feeling a bit stressed, aye.
>
> (Heterosexual women, public, Glasgow, focus group

One group of heterosexual men talked about how the inclusion of the HIV self-sampling kit could actually detract from the use of chlamydia self-sampling kits.

> Facilitator 2: Right, basically not do the tests?
>
> P3: Not do the tests. Because if I do the Chlamydia test and send it, where is the accompanying HIV test… which I, I’d hate to do. Facilitator 2: So basically, a lot more information and perhaps assurances for why…
>
> P3: Assurances for why it should be done, reasons why those tests need to be done; you can’t just find out about this and leave the other… because they’re kind of all in a group.
>
> P1: Ignorance is bliss. P3: Yeah. [Laughter]
>
> (Heterosexual men, public, London, focus group)

However, participants also talked of ways of minimising these barriers to Chlamydia self-sampling through making sure there was clarity concerning the choice of completing chlamydia self-sampling but not the HIV self-sample. Echoing the earlier findings, the sense of choice in selecting some, but not all, the self-sampling kits could be facilitated through attending to the structural elements of the pack itself.

#### Pack user return of samples for testing

The overall appearance of the sample return envelope and its contents raised some concerns for participants. These related primarily to the perceived stigma of STIs and other infectious diseases and concomitant perceptions that postal staff may not handle the return envelope correctly. Participants also outlined some barriers in relation to perceptions of the safety and effectiveness of the postal delivery system in enabling samples to get to the laboratory within the timeframe needed for samples to remain viable for testing. The extract below suggests a simple but effective solution to this perceived problem:

> P5: Do you get a notification when your tests arrive, your kit arrives at the… ? I think that would be my big, I know that you have to wait anyway but…
>
> P1: I never had one
>
> P5: That’s always been my concern about these things is like, I know that…
>
> P1: Has it got lost in the post? Has someone got a pot of my urine?
>
> P5: I know that’s potentially something that is almost, is it always going to be, is it two to four weeks, or whatever it is, you know, you don’t know but if it was anything that could Facilitator 2: Delivery receipt to say that it’s been - no, is that what you mean?
>
> P5: Yeah, yeah
>
> Facilitator 2: A delivery receipt to know that it’s gone to the right person.
>
> P5: That would be, even if it was just you know, when it gets to the lab, they send you a text.
>
> P2: Yeah
>
> (MSM, public, London, focus group)

Participants outlined how some of these concerns relating to the perceived efficacy of the postal system could be easily remedied through the provision of a range of return options, such as dropping off samples to a sexual health clinic, avoiding queues within clinics by using a ‘drop box’ facility where completed self-samples could be safely returned to a clinic to then be sent on to a lab. Participants acknowledged that the relative pros and cons of such an approach were moderated by the setting of the clinic, with those in more rural and small communities probably preferring postal routes rather than the expense, potential social exposure and felt stigma that might occur through using clinical contexts:

> P3: Conversely, I think for some people, the ability to just go and drop it in a post box is very appealing as well, as opposed to like if you live in a small town, you know, [NAME OF TOWN] if a bigger town, you..
>
> P1: Or having to wait in a queue or something like that, you know.
>
> P3: Yeah. The waiting room, of course, is full of your close friends, family and colleagues, as you drop off your sample, they’re all dropping off theirs.
>
> P4: Yeah, people in rural areas you’d imagine it could be very important.
>
> (Heterosexual men, public, Glasgow, focus group)

Table 1 (below) summarises the findings of the thematic analysis. The column on the left shows the key sequential step from Figure 2.

**Table 1:**
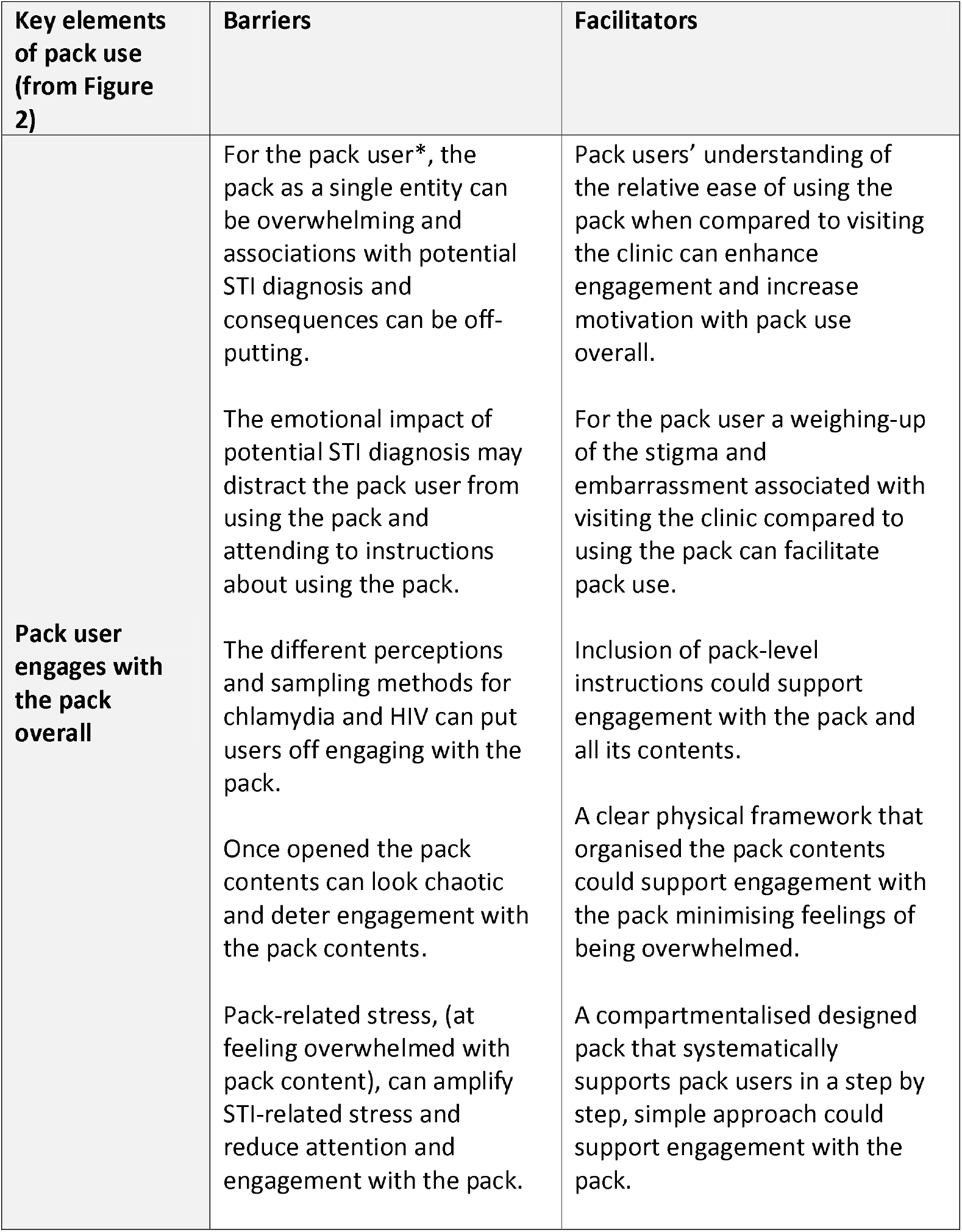
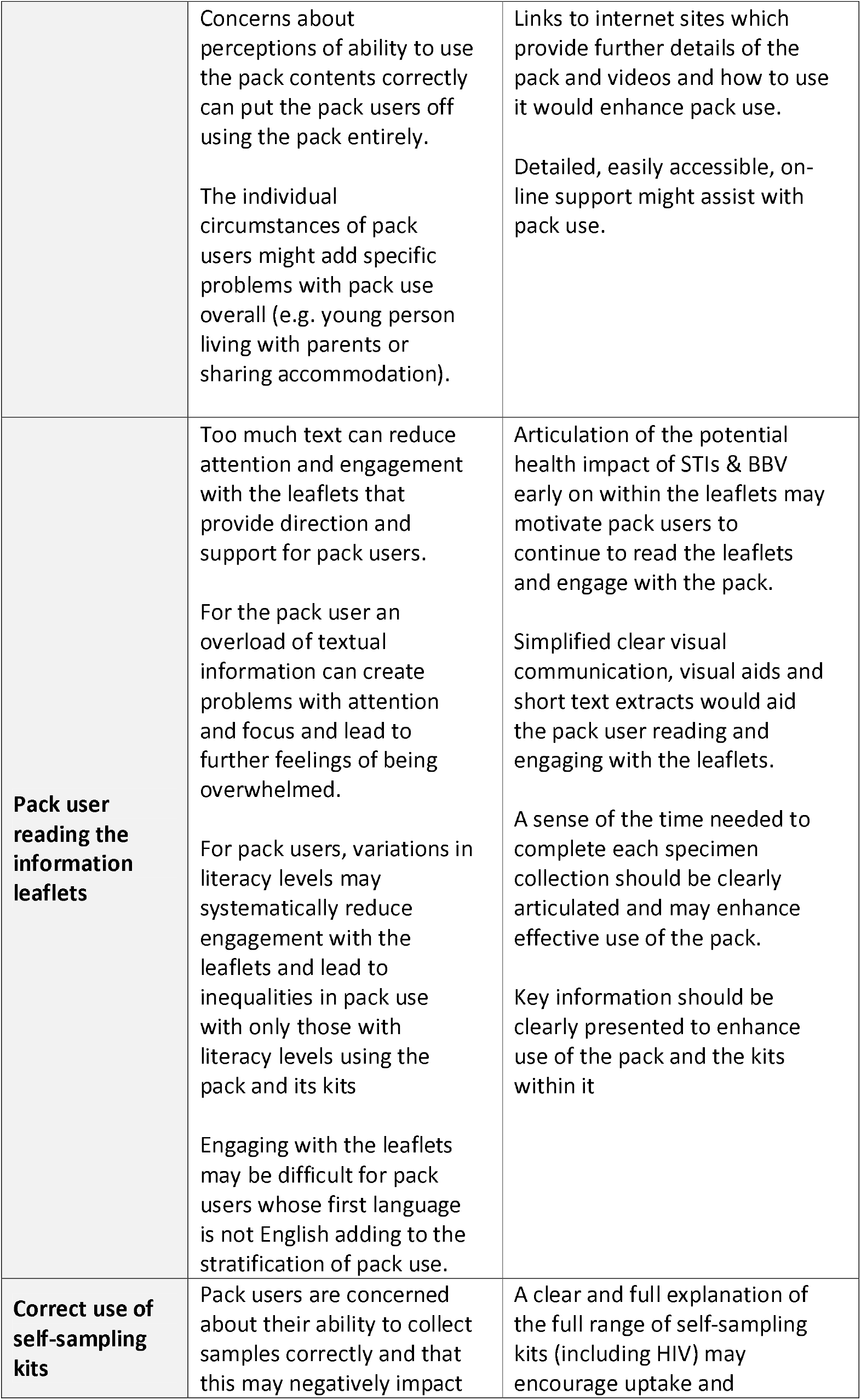
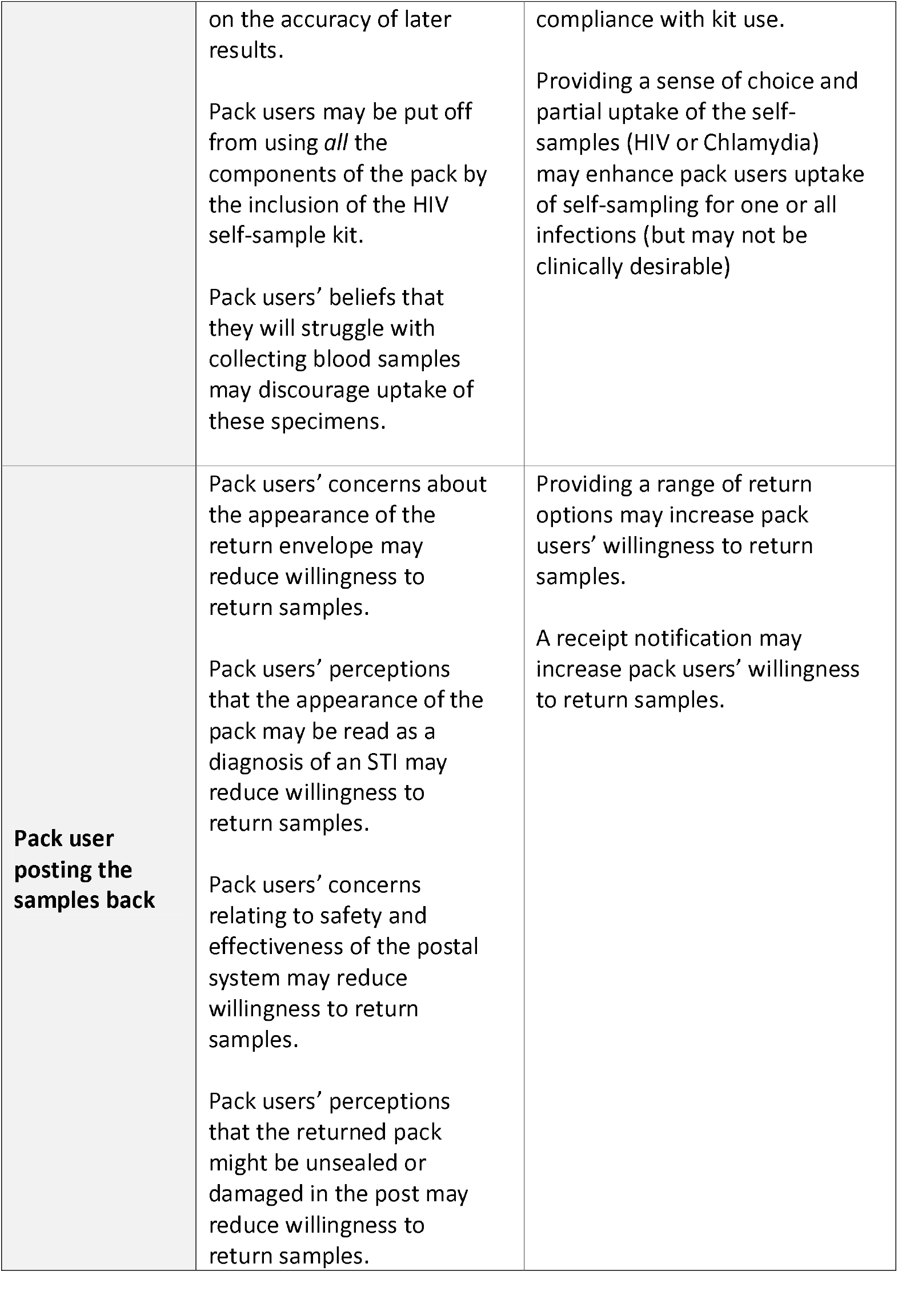
A summary of the key barriers and facilitators associated with use of the pack (key stages detailed in Figure 2)

## Discussion

Our thematic analysis of focus group data has delivered the first step in the process of intervention optimisation for a simple, pragmatic, intervention currently widely used across the UK (i.e. self-sampling packs for STIs & BBV). To our knowledge it is the first study to explore such packs and consider their optimisation in a systematic manner. Our paper is of particular interest to health psychologists as it focuses upon the use of qualitative data for an initial step in the process of intervention optimisation. Moreover, it is interesting as it includes a preliminary programme theory forming a key conceptual tool to be used iteratively throughout the wider intervention optimisation process, and indeed, future process evaluations. Although our data were collected and analysed before the COVID-19 pandemic it is interesting to anticipate how many of the issues explored here will also relate to COVID-19 self-sampling, or the self-sampling for a far wider range of health conditions that maybe instigated through social distancing.

Overall our analyses show that although in general these packs work well, there is considerable room for improvement. Our analysis shows the importance of engaging with the chain of sequential behaviours involved in pack use. Resolving issues early within the chain should have cascade effects on other behavioural elements enabling more people to use the pack and all of its contents. In other words it shows how initial consideration of the pack overall may be an important first step to widen access and increase use. The volume and organisation of pack contents was a barrier to some potential pack users. Equally, in relation to the content of the packs, our analysis highlighted particular problems with the instructions for the different self-sampling kits needed to test for a range of STIs and BBVs, these were too detailed and too wordy and could be improved with relatively simple adjustments to the formatting and presentation of kit instructions. In relation to findings regarding specific behaviours within our meaningful behavioural domains we found that within the domain of the ‘Correct use of self-sampling kits’ some of our participants differentiated the HIV self-sampling from the other self-sampling kits. Beyond this barrier being less important amongst MSM than heterosexuals, there was no particular socio-demographic pattern to identifying this behaviourally specific barrier. Finally, our analysis showed that there were some problems with the return of the samples that potentially could be removed with minor adjustments to the process.

Figure 3 illustrates how the findings reported here have implications for the development of our preliminary programme theory. We show how our findings (in red) suggest that psychological problems with the pack may well have medium- and long-term impacts that amplify existing health inequalities. Again, although not the focus of the current study it is interesting to consider how these same dynamics may be important for self-sampling for COVID-19.

**Figure 3:**
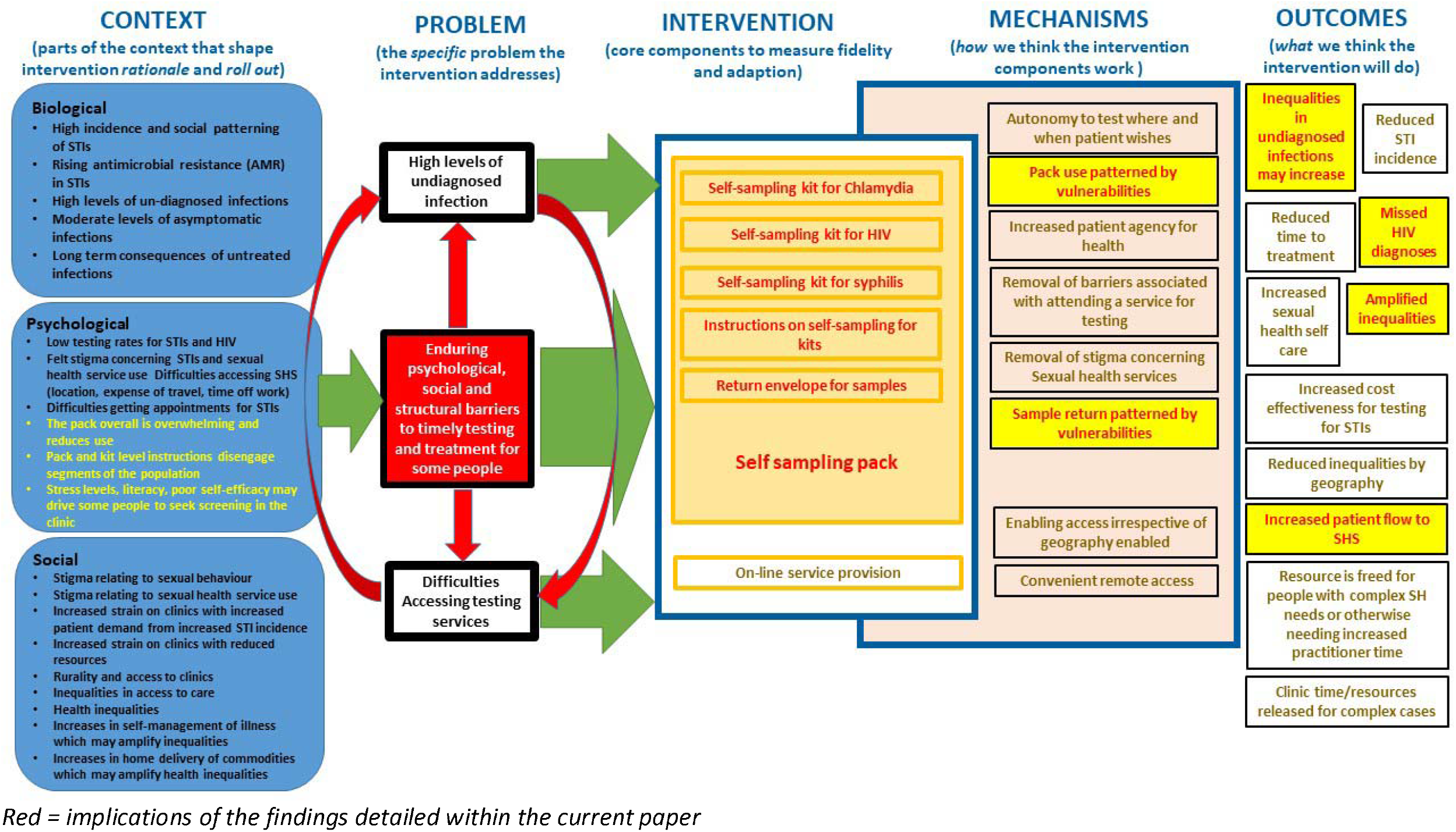
Revised programme theory illustrating the results of thematic analyses.

Our approach to understanding the behavioural system of self-sampling pack use and the range of interconnected behavioural domains is vital for executing the subsequent steps of intervention optimisation effectively. We would suggest that current guidance on how to select behaviours to target within intervention development often underestimates the complexity and interdependencies of the elements of behavioural systems (Michie et al., 2014). Within the current project, and its focus on intervention optimisation, we sought to have a distinct inductive analytic focus on understanding the behavioural aspects of the use of self-sampling packs from the pack user’s perspective. Overall our approach to pragmatically identifying meaningful behavioural domains worked well, with the exception of the participant-driven distinction between the HIV and other STI sampling kits. This step of conceptualising the behavioural foci was necessary to facilitate the more deductive analyses matching appropriate barriers and facilitators to specific behavioural domains and concomitant behaviours. In turn this was essential to enable the delivery of the subsequent behaviour change wheel analyses that followed (Flowers *et al*., forthcoming 2020).

The analysis here is limited by its sole reliance on qualitative methods. If it had been possible within the budget and programme timeline, we would have complemented our qualitative assessment of barriers and facilitators to pack use reported here with additional quantitative assessment. Furthermore, the bulk of the analysis was conducted by one experienced analyst rather than coded by a wider group of researchers within the team. Along these lines it is hard to appraise the relative importance of each barrier or facilitator in any objective manner. However, at a later stage within the whole intervention optimisation process we used the APEASE criteria (Michie et al., 2014) with a group of mixed health professionals to address eventual recommendations in terms of Acceptability, Practicability, Effectiveness/cost-effectiveness, Affordability, Safety/side-effects and Equity. This analysis is reported elsewhere (Flowers *et al*., forthcoming 2020). In this way the overall project provided a number of safety checks to reduce the risks associated with our approach. In addition, the context in which our data were collected potentially shapes the value of our findings. We collected the data as part of a wider intervention optimisation process of a partner notification intervention (Estcourt et al., 2020). As such the data were collected in the context of a study where participants were primed by our research procedures to consider situations in which people who used the packs were at a high chance of having acquired an STI. Moreover when packs were discussed the packs included antibiotic treatment for STIs.

## Conclusion

This study provides an exemplar of how to use qualitative data generated through focus groups and both inductive and deductive thematic analysis to understand the factors shaping a series of inter-related behavioural domains concerning a widely used complex intervention intended to improve sexual health (e.g., packs for sexual health and blood borne virus self-sampling). The paper provides an account of an initial step within a larger process of intervention optimisation in the absence of a prior framework made explicit in an intervention development phase. We provide empirical evidence of both the overall behavioural system of pack use, and the barriers and facilitators to the meaningful behavioural domains within that behavioural system. In addition we present and iteratively use an initial programme theory to show how our work contributes to understanding and situating the behaviour change problem at the heart of the optimisation of the self-sampling pack.

## Data Availability

N/A

## Acknowledgements

The authors would like to thank all of the participants who took part in the focus groups and interviews and all individuals and organisations who helped with recruitment for this study via social media, word of mouth and direct contact to their networks.

This study has been shaped through ongoing discussion and support from the whole LUSTRUM team (Claudia S Estcourt (Principal Investigator), Alex Comer, Alison R Howarth, Andrew Copas, Anna Tostevin, Anne M Johnson, Catherine H Mercer, Chidubem (Duby) Ogwulu, Christian Althaus, Fiona Mapp, Gabriele Vojt, Jackie Cassell, Jean McQueen, John Saunders, Karen Pickering, Maria Pothoulaki, Melvina Woode Owusu, Merle Symonds, Nicola Low, Oliver Stirrup, Paul Flowers, Rak Nandwani, Sonali Wayal, Susannah Brice, Tracy Roberts) and the Programme Steering Committee (PSC) (Simon Barton (chair), Alex Miners, Artemis Koukounari, David Crundwell, Emmanuel Rollings-Kamara, Lynis Lewis, Rebecca Turner, Robbie Currie, Rachel Shaw, Saima Saddiqui (on behalf of NIHR programme management).

## Contributor statement

CSE, PF, JAC and JS conceptualized the LUSTRUM programme including the design of this study and acquired funding. MP, GV, FM and MWO collected the data. PF, MP, and GV analysed the data. PF wrote the first draft. All authors commented on paper drafts and contributed to the editing and reworking of the paper. All authors gave approval of the final version for submission.

## Ethical approval

Ethical approval from Glasgow Caledonian University Research Ethics Committee (HLS/PSWAHS/A15/256) and NHS Ethics Approval (16/NI/0211) were obtained.

## Disclaimer

The views expressed are those of the author(s) and not necessarily those of the NHS, the NIHR or the Department of Health. The funders had no role in study design, collection, management, analysis and interpretation of data; writing of the report and the decision to submit the report for publication.

## References

Banerjee, P., Thorley, N. and Radcliffe, K. (2018) ‘A service evaluation comparing home-based testing to clinic-based testing for Chlamydia and gonorrhoea in Birmingham and Solihull’, International Journal of STD and AIDS. SAGE Publications Ltd, 29(10), pp. 974–979. doi: 10.1177/0956462418767180.

Barnard, S. et al. (2018) ‘Comparing the characteristics of users of an online service for STI self-sampling with clinic service users: A cross-sectional analysis’, Sexually Transmitted Infections. BMJ Publishing Group, 94(5), pp. 377–383. doi: 10.1136/sextrans-2017-053302.

Brousselle, A. and Champagne, F. (2011) ‘Program theory evaluation: Logic analysis’, Evaluation and Program Planning, 34(1), pp. 69–78. doi: 10.1016/j.evalprogplan.2010.04.001.

Campbell, D. and Marsh, S. (2017) STI warning as clinics close in London and self-testing is delayed, The Guardian. Available at: https://www.theguardian.com/society/2017/oct/15/sti-warning-as-clinics-close-in-london-and-self-testing-is-delayed (Accessed: 13 August 2020).

Cowan, F. M., French, R. and Johnson, A. M. (1996) ‘The role and effectiveness of partner notification in STD control: a review.’, Genitourinary medicine. BMJ Publishing Group, 72(4), pp. 247–52. Available at: http://www.ncbi.nlm.nih.gov/pubmed/8976827 (Accessed: 31 May 2016).

Engel, G. L. (1977) ‘The Need for a New Medical Model: A Challenge for Biomedicine’, Science, 196(4286), pp. 129–36.

Estcourt, C. et al. (2012) ‘Can we improve partner notification rates through expedited partner therapy in the UK? Findings from an exploratory trial of Accelerated Partner Therapy (APT)’, Sexually Transmitted Infections, 88(1), pp. 21– 26. doi: 10.1136/sti.2010.047258.

Estcourt, C. S. et al. (2015) ‘Developing and testing accelerated partner therapy for partner notification for people with genital Chlamydia trachomatis diagnosed in primary care: A pilot randomised controlled trial’, Sexually Transmitted Infections. BMJ Publishing Group Ltd, 91(8), pp. 548–554. doi: 10.1136/sextrans-2014-051994.

Estcourt, C. S. et al. (2020) ‘Accelerated partner therapy (APT) partner notification for people with Chlamydia trachomatis: Protocol for the Limiting Undetected Sexually Transmitted infections to RedUce Morbidity (LUSTRUM) APT cross-over cluster randomised controlled trial’, BMJ Open, 10(3), p. 34806. doi: 10.1136/bmjopen-2019-034806.

Flowers, P. et al. (2020) Using the behaviour change wheel approach to help optimise home-based self-sampling packs for sexually transmitted infections and blood borne viruses.

Guerra, L. et al. (2016) ‘National HIV self-sampling service’, in National HIV Prevention England Conference. London, UK: Public Health England. Available at: https://www.hivpreventionengland.org.uk/wp-content/uploads/2017/06/Luis-Guerra-_National-HIV-Home-Sampling-Service-1.pdf.

Kersaudy-Rahib, D. et al. (2017) ‘Chlamyweb Study II: A randomised controlled trial (RCT) of an online offer of home-based Chlamydia trachomatis sampling in France’, Sexually Transmitted Infections, 93(3), pp. 188–195. doi: 10.1136/sextrans-2015-052510.

Manavi, K. and Hodson, J. (2017) ‘Observational study of factors associated with return of home sampling kits for sexually transmitted infections requested online in the UK’, BMJ Open. BMJ Publishing Group, 7(10), p. e017978. doi: 10.1136/bmjopen-2017-017978.

Medical Research Council (no date) Developing and evaluating complex interventions.

Michie, S., Atkins, L. and West, R. (2014) The Behaviour Change Wheel: A Guide To Designing Interventions. 1st ed. London, UK: Silverback Publishing.

Michie, S., van Stralen, M. M. and West, R. (2011) ‘The behaviour change wheel: A new method for characterising and designing behaviour change interventions’, Implementation Science, 6(1). doi: 10.1186/1748-5908-6-42.

Morris, J. L. et al. (2014) ‘Sexually transmitted infection related stigma and shame among African American Male Youth: Implications for testing practices, partner notification, and treatment’, AIDS Patient Care and STDs, 28(9), pp. 499–506. doi: 10.1089/apc.2013.0316.

Pothoulaki, M. et al. (in preparation) Intervention development and specification for the optimal delivery of APT.

Rogers, P. J. (2008) ‘Using Programme Theory to Evaluate Complicated and Complex Aspects of Interventions’, Evaluation, 14(1), pp. 29–48. doi: 10.1177/1356389007084674.

Tanton, C. et al. (2017) ‘Sexual health clinic attendance and non-attendance in Britain: findings from the third National Survey of Sexual Attitudes and Lifestyles (Natsal-3).’, Sexually transmitted infections. doi: 10.1136/sextrans-2017-053193.

White, C. (2017) ‘Sexual health services on the brink’, BMJ (Clinical research ed.). NLM (Medline), 359, p. j5395. doi: 10.1136/bmj.j5395.

Woodhall, S. C. et al. (2016) ‘Is chlamydia screening and testing in Britain reaching young adults at risk of infection? Findings from the third National Survey of Sexual Attitudes and Lifestyles (Natsal-3)’, Sexually Trans Infect, 92, pp. 218–227. doi: 10.1136/sextrans-2015-052013.

World Health Organization (2018) Report on global sexually transmitted infection surveillance 2018. Geneva: World Health Organization. Available at: http://www.who.int/reproductivehealth/publications/stis-surveillance-2018/en/ (Accessed: 12 August 2020).

